# Anagliptin monotherapy in patients with type 2 diabetes mellitus and high low-density lipoprotein cholesterol reduces fasting plasma lathosterol level: a single-arm intervention trial

**DOI:** 10.1101/2020.05.16.20095307

**Authors:** Yuichi Ikegami, Ikuo Inoue, Yasuhiro Takenaka, Daigo Saito, Mitsuhiko Noda, Akira Shimada

**Affiliations:** Department of Endocrinology and Diabetes, Saitama Medical University, Faculty of Medicine, 38 Morohongo, Moroyamacho, Irumagun, Saitama 350-0495, Japan; Department of Physiology, Graduate School of Medicine, Nippon Medical School, 1-25-16 Nezu, Bunkyo, Tokyo 113-0031, Japan

**Keywords:** anagliptin, dipeptidyl peptidase-4 inhibitor, low-density lipoprotein cholesterol, lathosterol, glucagon-like peptide-1

## Abstract

**Background:** Anagliptin, a dipeptidyl peptidase-4 (DPP-4) inhibitor, has been shown to decrease low-density lipoprotein cholesterol (LDL-C) levels in plasma.

**Aim of study:** The objective of our study is to elucidate the mechanisms responsible for anagliptin-mediated improvements in high LDL-C levels (hyper-LDL-C-emia).

**Methods:** We prospectively examined the effects of anagliptin monotherapy on fasting plasma lathosterol, sitosterol, and campesterol levels in patients with type 2 diabetes mellitus and hyper-LDL-C-emia for 6 months. We examined 8 patients who did not use hypoglycemic or lipid-lowering drugs, other than anagliptin, for 4 months before initiating the study. Serum variables related to glucose and lipid metabolism were measured before and after the treatment for 6 months and pre- and post-prandially using the cookie-loading test.

**Results:** After treatment, anagliptin monotherapy (n = 8) significantly decreased fasting LDL-C (182.8 to 167.8 mg/dL, as mean values of before and after the treatment) and plasma lathosterol levels (3.2 to 2.6 mg/dL); however, no significant changes were observed in fasting sitosterol or campesterol levels. Furthermore, a significant increase (*p* = 0.0012) in the change in 1-h post-prandial active glucagon-like peptide-1 (GLP-1) levels was observed after anagliptin treatment. For all participants (n = 17), fasting plasma lathosterol levels were negatively correlated with pre-prandial GLP-1 levels.

**Conclusion:** Anagliptin monotherapy may have a beneficial effect on lipid metabolism, which is mediated by the inhibition of hepatic cholesterol synthesis, and not by the inhibition of intestinal lipid transport.

## 1. Introduction

Dipeptidyl peptidase-4 (DPP-4) inhibitors increase the plasma levels of incretins, such as glucagon-like peptide-1 (GLP-1), by selectively inhibiting DPP-4 and thereby promoting insulin secretion, which then results in hypoglycemic effects in patients with type 2 diabetes mellitus (T2DM). The degree of insulin secreted by the pancreatic beta cells, following the stimulation of incretins, is dependent on blood glucose levels. Therefore, the risk of hypoglycemia by DPP-4 inhibitor treatments is lower than that by other diabetes drugs, such as sulfonylureas (SUs). DPP-4 inhibitors are currently used worldwide, and each DPP-4 inhibitor has different clinical features depending on the type of agent [1]. Similar to other DPP-4 inhibitors, anagliptin decreases glycosylated hemoglobin (HbA1c) and plasma glucose levels [2, 3]. It has been shown to achieve the strongest effect in increasing plasma GLP-1 levels by administering a dose of 100-200 mg twice daily to patients with T2DM [4]. A previous study has reported that anagliptin treatment decreases plasma low-density lipoprotein cholesterol (LDL-C) levels [5]. Thus, in the present study, we examined the effects of anagliptin on fasting plasma lathosterol, sitosterol, and campesterol levels in patients with T2DM and hyper-LDL-C-emia for 6 months to elucidate the mechanisms that lead to improvements in plasma LDL-C levels. Lathosterol is a whole-body cholesterol synthesis marker in the liver, while sitosterol and campesterol (both plant sterols) are sterol absorption markers in the intestines.

## 2. Materials and Methods

### 2.1 Subjects

The inclusion criterion of the present study was as follows: patients with poorly controlled T2DM (HbA1c < 8.0% or 63mmol/mol) and an LDL-C level of more than 140 mg/dl. Participating T2DM patients had inadequate glycemic and lipid control and were being treated with dietary and exercise interventions. The participants aged between 20 and 80 years of males and females. Key exclusion criteria were as follows: 1) patients using hypoglycemic agents (SUs, biguanides, alpha-glucosidase inhibitors, glinides, dipeptidyl peptidase-4 inhibitors, insulin, and GLP-1 receptor agonists); 2) hypersensitivity to anagliptin; 3) patients with severe diabetic ketoacidosis, diabetic coma/pre-coma, or type 1 diabetes; 4) patients with sepsis, those scheduled for surgery and who underwent surgery, and those with severe trauma; 5) patients with severe renal dysfunction (serum creatinine level > 2.4 mg/dl for male and > 2.0 mg/dl for female), and patients undergoing hemodialysis or peritoneal dialysis; 6) female patients who were pregnant or suspected to be pregnant; and 7) patients deemed by the investigator to be ineligible for participation in the study.

### 2.2. Study Protocol

A dose of 100-200 mg of anagliptin was administered in the morning and in the evening. The Institutional Review Board of Saitama Medical University Hospital approved the experimental protocol. This was a single arm, non-randomized, open-label prospective study with blinded assessors. The protocol was registered at the University Hospital Medical Information Network (UMIN) on August 5, 2013, with the identification number UMIN000011256. Patients who provided written informed consent were enrolled in the pre-drug intervention phase for 4 months with only lifestyle interventions; this was done to ensure that any observed effects with pharmacotherapy were attribute to anagliptin monotherapy. We assessed the following parameters at the start of the pre-drug period: both pre- and 1- and 2-h post-prandial blood levels of glucose; lipid, insulin, C-peptide, and proinsulin levels; pre- and 1-h post-prandial active GLP-1 levels using the cookie-loading test (CLT); and fasting plasma lathosterol, sitosterol, and campesterol levels. During this period, 6 patients dropped out because of the necessity for interventions other than anagliptin for dyslipidemia or hyperglycemia. After the pre-drug intervention phase, patients received anagliptin monotherapy for 6 months. During the last visit of this study, patients underwent CLT again. To assess the effects of anagliptin monotherapy on lipid metabolism, patients receiving drugs other than anagliptin were excluded from the main analysis.

### 2.3. Measurements

The metabolic parameters described above, including glucose and lipid levels, were measured at SRL, Inc. (Tokyo, Japan). These data were obtained to investigate the effects of 6-month anagliptin monotherapy on both pre- and 1- and 2-h post-prandial levels of blood glucose, lipid, insulin using Lumipulse® PrestoInsulin (Fujirebio Inc., Tokyo, Japan), C-peptide using Lumipulse® PrestoC-peptide (Fujirebio Inc., Tokyo, Japan), proinsulin using Human Proinsulin RIA KIT (Millipore Cor., MA, USA), and pre- and 1-h post-prandial active GLP-1 levels using Glucagon-Like Peptide-1 (Active) ELISA KIT (Millipore Cor., MA, USA) in participating patients after 6 months. Moreover, markers for cholesterol synthesis (lathosterol) and absorption (sitosterol and campesterol) were determined using gas chromatography, GC-2010 (Shimadzu Co., Ltd, Kyoto, Japan). Serum apoB-48 levels were measured using Apo B-48 chemiluminescent enzyme immunoassay KIT (Fujirebio Inc., Tokyo, Japan); these levels reflect exogenous lipoproteins, such as chylomicrons, and their remnants. LDL-C levels were calculated using the Friedewald equation because all patients had triglyceride (TG) levels less than 400 mg/dl.

### 2.4. CLT

CLT was performed after overnight fasting using previously described methods at baseline and at 6 months after starting anagliptin treatment. Patients were instructed to ingest a cookie with water within 30 minutes: half of the cookie was to be eaten within 10 min and the remainder within the last 20 min. Anagliptin was administered immediately after CLT. Time measurements were started when half the cookie was ingested. Blood samples were obtained at 0, 1, and 2 h after ingesting the cookie, which consisted of 75 g of carbohydrates, 25 g of fat, and 8 g of protein, and 592 kcal of energy (Saraya Co., Ltd., Osaka, Japan).

### 2.5. Data analysis

The primary outcome was the change in LDL-cholesterol levels after 6 months. The secondary outcome was changes in plasma lathosterol, sitosterol, and campesterol levels. Data are presented as the mean ± standard deviation (SD) of the parametric variables. Differences before and after anagliptin treatment were examined using the paired Student’s *t*-test, and the significance was set at a *p*-value of less than 0.05. A sub-analysis of the correlation between lathosterol and active GLP-1 levels was performed by Spearman’s correlation test using samples from the remaining 17 participating patients at the end of the study. All analyses were performed using the Statistical Package for Social Association Version 21.0 (SPSS Inc., Chicago, IL)

## 3. Results

Twenty-three patients with T2DM and hyper-LDL-C-emia were included in the study. Both pre- and 1- and 2-h post-prandial levels of blood glucose, lipids, insulin, C-peptide, and proinsulin; pre- and 1-h post-prandial active GLP-1 levels; and fasting plasma lathosterol, sitosterol, and campesterol levels were measured. Six patients dropped out because of worsening hyperlipidemia and/or hyperglycemia before the initiation of the anagliptin treatment. Among the 17 patients who were followed until the end of the study, 1 patient received additional treatment with 10-mg ezetimibe daily and 2 patients received 5-mg pravastatin daily to manage hyperlipidemia. We mainly analyzed 8 patients for whom therapeutic agents were not changed during the test period (Fig. 1). These 8 patients did not use hypoglycemic agents, such as SUs, biguanides, alpha-glucosidase inhibitors, glinides, thiazolidinedione, other DPP-4 inhibitors, sodium-glucose co-transporter 2 inhibitors, insulin, and GLP-1 receptor agonists, and did not use oral drugs for hyperlipidemia including statins, fibrates, and ezetimibe.

**Figure 1.**
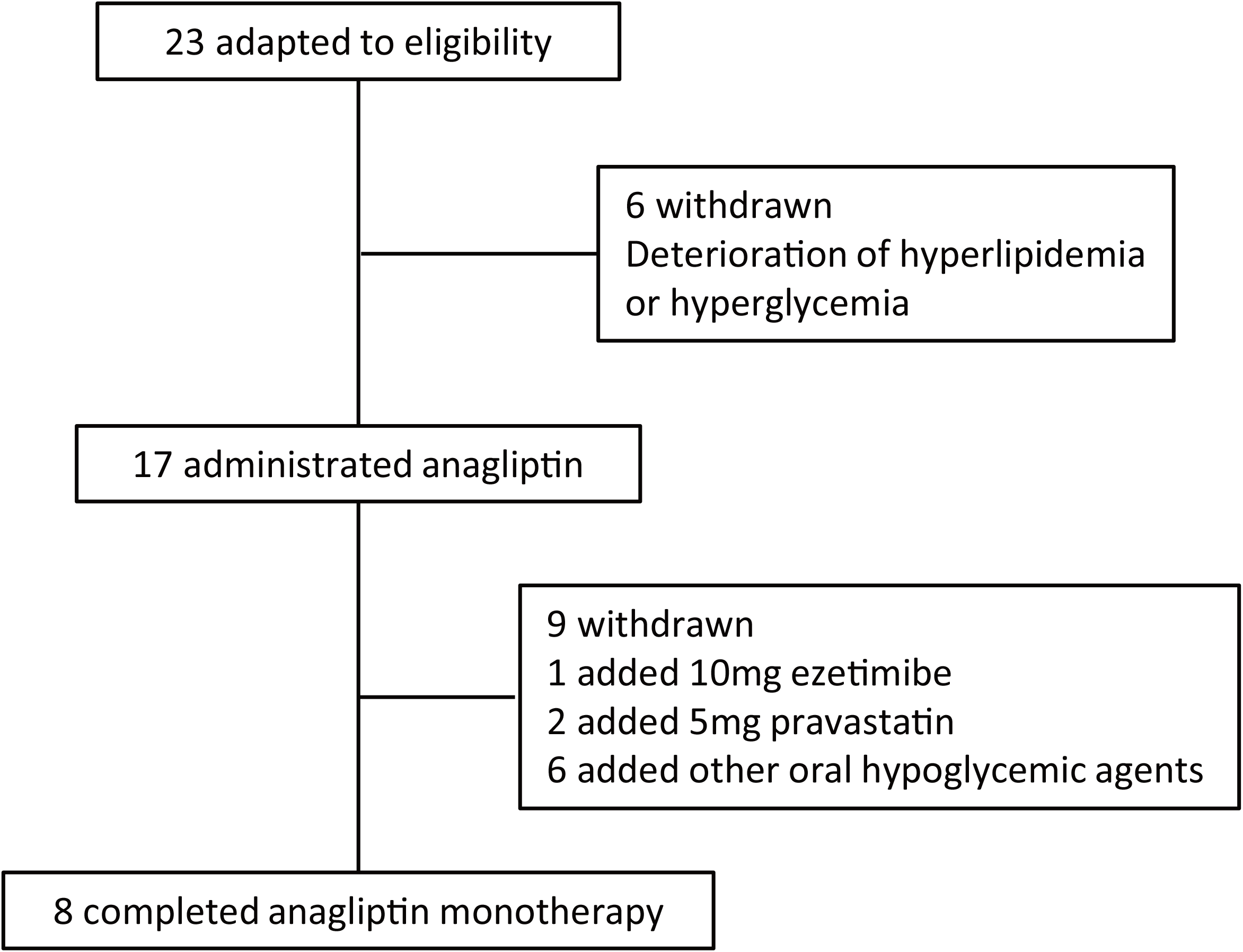
Flowchart of participant selection in this study.

The demographics of these 8 patients are summarized in Table 1, and their biochemical characteristics before and after anagliptin monotherapy are shown in Table 2. After 6 months of anagliptin monotherapy, HbA1c levels significantly decreased in these 8 patients. Anagliptin monotherapy for 6 months significantly decreased fasting lathosterol levels (*p* < 0.05), whereas no such significant changes were noted in fasting sitosterol or campesterol levels (Table 2).

**Table 1.** Baseline demographics of eight patients receiving anagliptin monotherapy. Data on age and body mass index are expressed as means ± standard deviations.

| Demographics (n=8) |  |
| --- | --- |
| Female sex-no. of patients (%) | 4 (50) |
| Age-year | 68.1±6.7 |
| Body mass index-kg/m<sup>2</sup> | 24.6±4.2 |

**Table 2.** Comparison of parameters of eight patients before and after treatment with anagliptin monotherapy. Data are expressed as means ± standard deviations, and are comparisons of changes between baseline and at 6 months of treatment for eight patients. ^a^We used detection-limited value (3.1 pmol/L) for one patient after 6 months of treatment. BMI, body mass index; FPG, fasting plasma glucose; TC, total cholesterol; LDL-C, low-density lipoprotein cholesterol; HDL-C, high-density lipoprotein cholesterol; TG, triglyceride. The conversion factor was as follows: glucose (mmol/L) = glucose (mg/dL)/18.0; cholesterol (mmol/L) = cholesterol (mg/dL)/38.6; triglyceride (mmol/L) = triglyceride (mg/dL)/88.5; insulin (pmol/L) = insulin (μIU/mL)/0.144.

| Biochemical characteristics (n=8) |  |  |  |
| --- | --- | --- | --- |
|  | Baseline | 6 months | *p* |
| BMI (kg/m<sup>2</sup>) | 24.6±3.9 | 24.8±4.4 | 0.5325 |
| FPG (mg/dL) | 129.8±13.5 | 120.6±9.8 | 0.1475 |
| HbA1c (%) | 6.5±0.3 | 6.1±0.1 | 0.0033 |
| TC (mg/dL) | 260.4±42.6 | 246.9±45.3 | 0.0306 |
| LDL-C (mg/dL) | 182.8±34.7 | 167.8±38.3 | 0.0159 |
| HDL-C(mg/dL) | 58.0±10.6 | 56.0±10.5 | 0.1819 |
| TG (mg/dL) | 97.8±30.8 | 115.3±44.8 | 0.1179 |
| Insulin (μU/mL) | 7.6±3.6 | 8.1±5.5 | 0.6530 |
| Proinsulin (pmol/L) <sup>a</sup> | 20.2±14.6 | 18.9±13.9 | 0.2733 |
| Lathosterol (μg/mL) | 3.2±1.0 | 2.6±0.5 | 0.0413 |
| Sitosterol (μg/mL) | 3.1±1.1 | 3.1±1.1 | 1.0000 |
| Campesterol (μg/mL) | 5.6±1.6 | 5.8±1.8 | 0.3459 |

Plasma glucose, insulin, and proinsulin levels obtained before and after anagliptin monotherapy using the CLT are shown in Figure 2. Plasma glucose and proinsulin levels significantly decreased after 6 months of anagliptin monotherapy, whereas no significant changes were noted in insulin levels. After 6 months of anagliptin monotherapy, glucose levels at 1 and 2 h after the CLT reduced significantly; however, no significant changes were noted in fasting glucose levels. Plasma insulin levels before and 1 and 2 h after the CLT did not significantly increase by 6-months anagliptin monotherapy. However, proinsulin levels were significantly decreased 2 h after the CLT. Plasma GLP-1 levels before and 1 h after the CLT at the end of anagliptin monotherapy are shown in Figure 3. Post-prandial plasma GLP-1 levels increased significantly following anagliptin monotherapy. After 6 months, plasma TC and high-density lipoprotein cholesterol (HDL-C) levels before and after the CLT were not significantly different. However, plasma TG and apoB-48 levels slightly increased (Fig. 4). The value of LDL-C was calculated using the Friedewald method (from plasma TC, TG, and HDL-C levels). After 6 months of anagliptin monotherapy, plasma LDL-C levels before the CLT decreased significantly, but not at 1 or 2 h after the CLT (Fig. 5).

**Figure 2.**
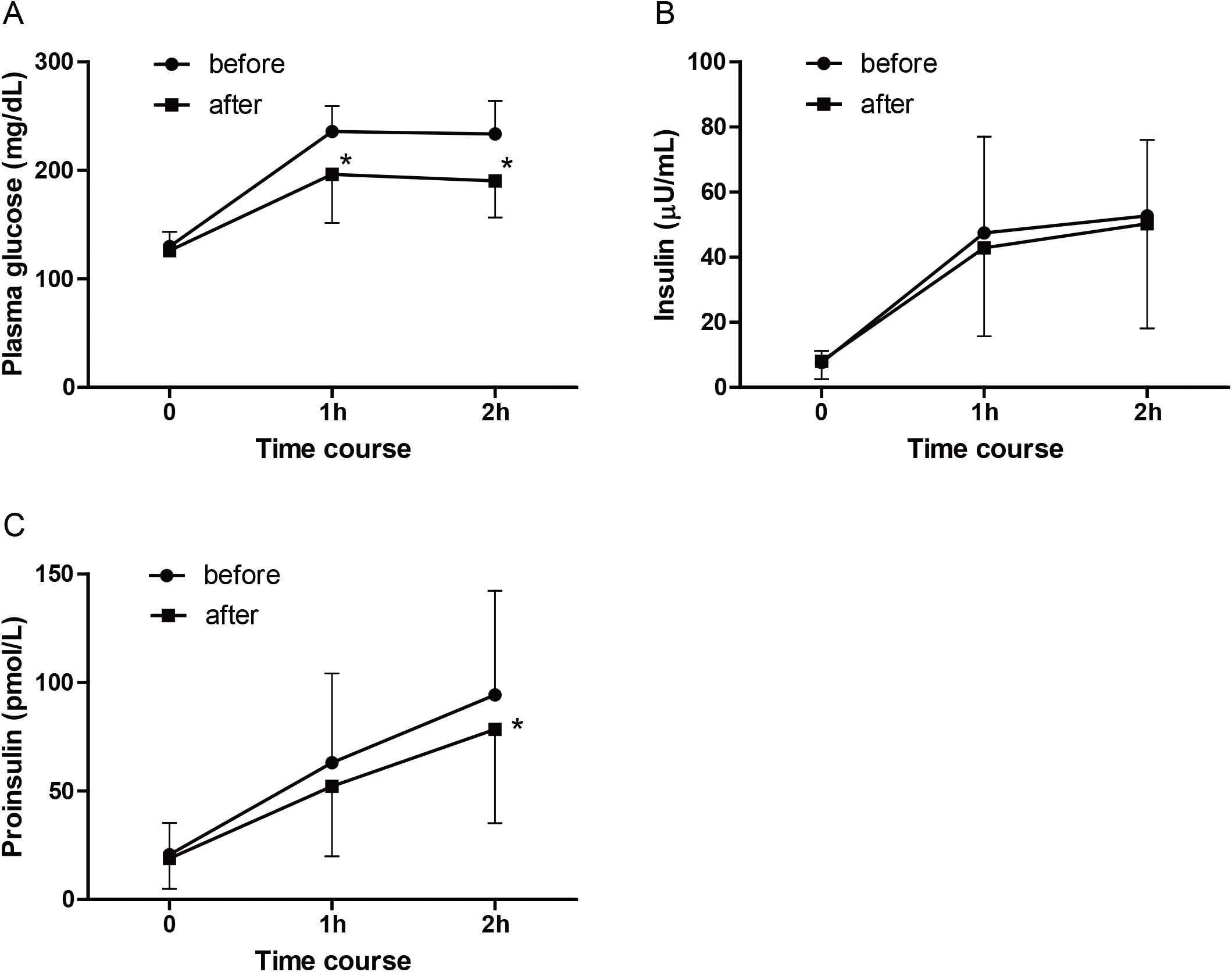
Changes in parameters assessed using cookie-loading test. Plasma glucose (A), insulin (B), and proinsulin(C) levels. The conversion factor was as follows: glucose (mmol/L) = glucose (mg/dL)/18.0; insulin (pmol/L) = insulin (μIU/mL)/0.144. *\*p* < 0.05 versus before treatment.

**Figure 3.**
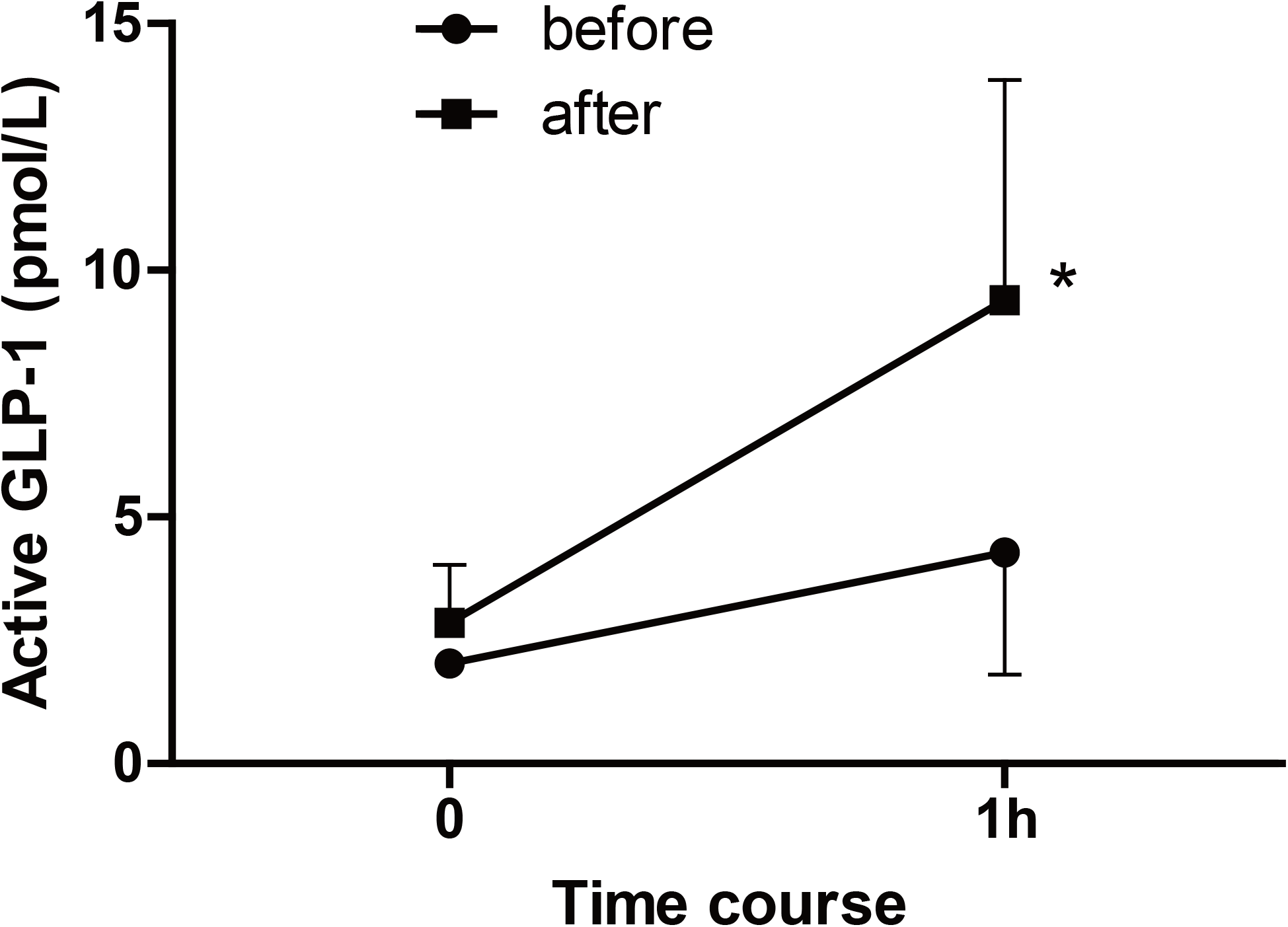
Assessing changes in active glucagon-like peptide-1 using cookie-loading test. *\*p* < 0.05 versus before treatment.

**Figure 4.**
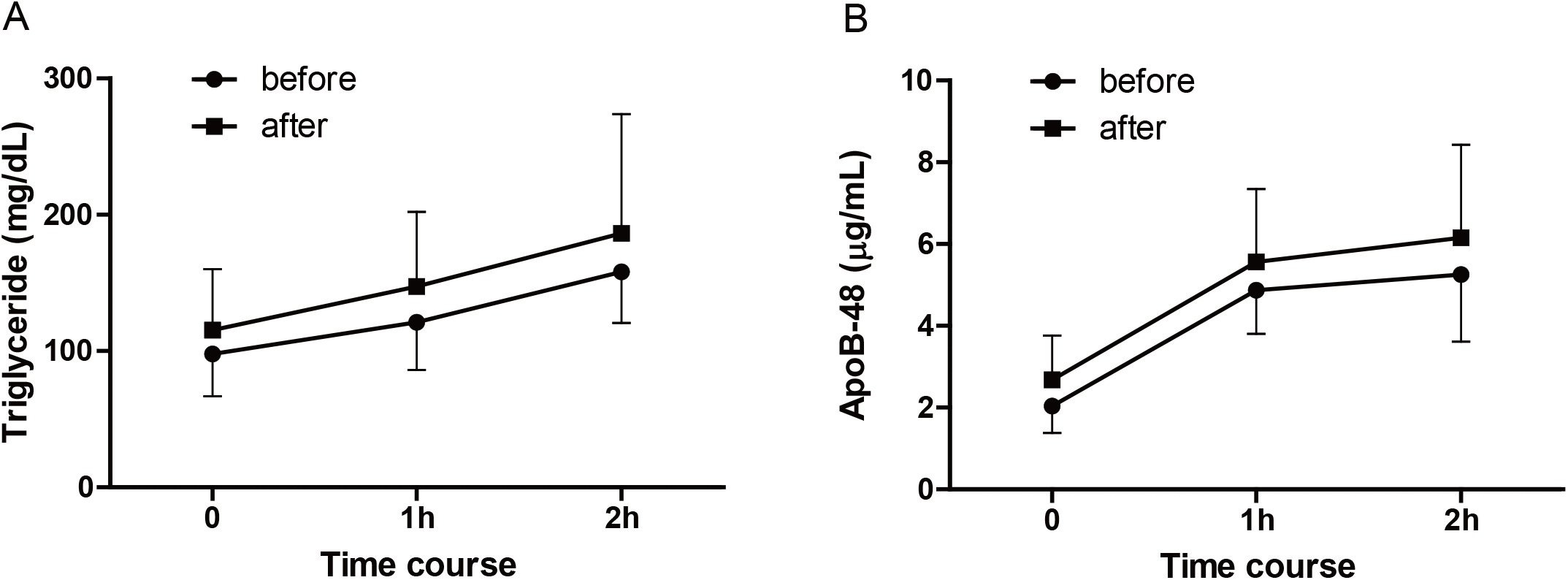
Assessing changes in parameters using cookie-loading test. Triglyceride (A) and apoB-48 (B) levels. The conversion factor was as follows: triglyceride (mmol/L) = triglyceride (mg/dL)/88.5.

**Figure 5.**
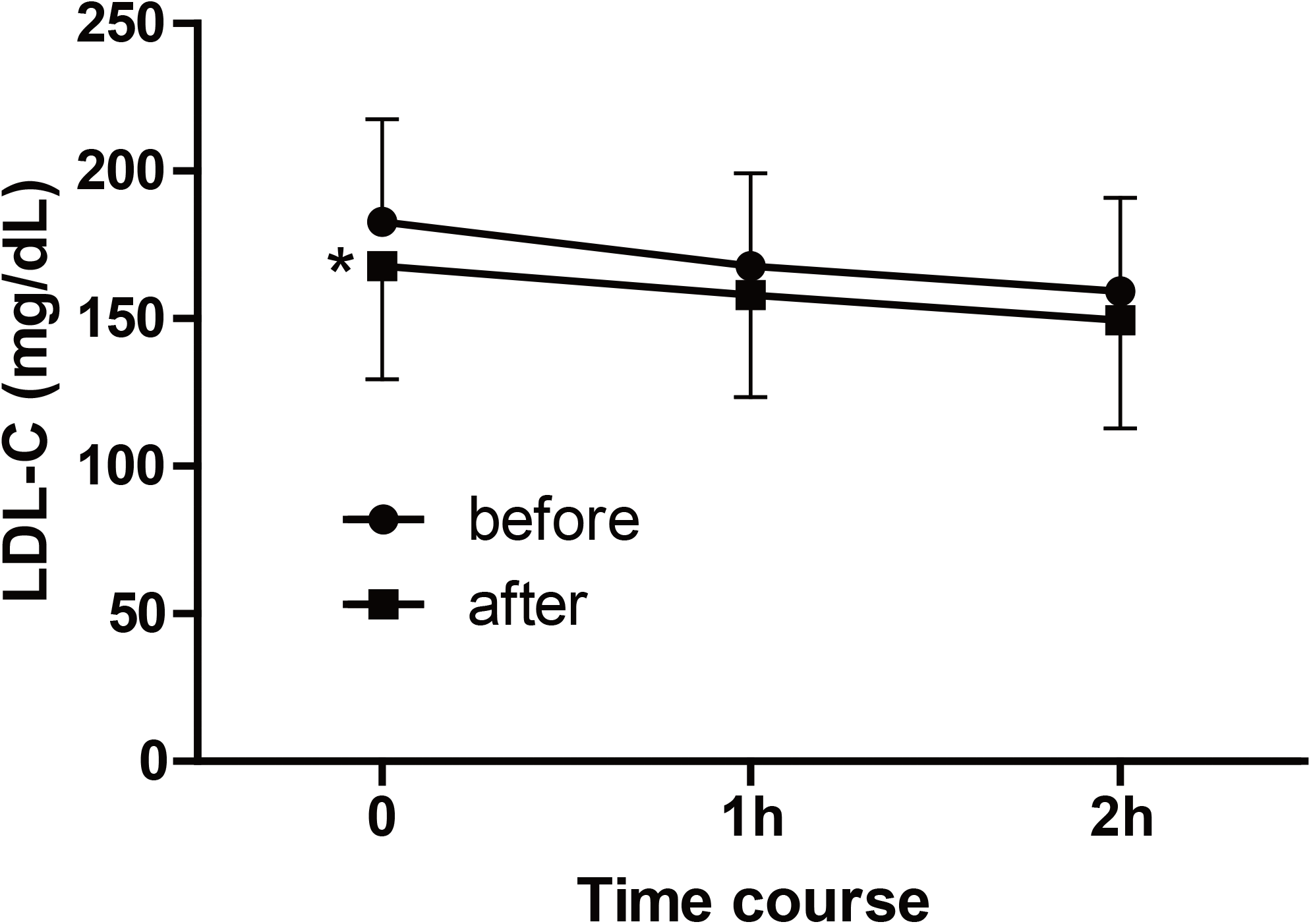
Assessing changes in low-density lipoprotein cholesterol using cookie-loading test. The conversion factor was as follows: cholesterol (mmol/L) = cholesterol (mg/dL)/38.6. *\*p* < 0.05 versus before treatment.

We examined the relationship between active GLP-1 and lathosterol, campesterol, or sitosterol levels as a sub-analysis. Figure 6A shows a scatter plot of the relationship between fasting active GLP-1 levels and fasting plasma lathosterol levels in all 17 patients at end of the study. Since GLP-1 levels of 2.0 pmol/L or lower were not detectable, plots corresponding to these levels were omitted from the figure. Similarly, Figures 6B and 6C show scatter plots corresponding to the relationship between fasting active GLP-1 levels and fasting plasma campesterol or sitosterol levels. A negative correlation was observed between fasting active GLP-1 levels and fasting plasma lathosterol levels but not between the former and fasting plasma campesterol or sitosterol levels. No correlation was noted between post-prandial active GLP-1 levels and fasting plasma lathosterol, campesterol, and sitosterol levels at the CLT (data not shown).

**Figure 6.**
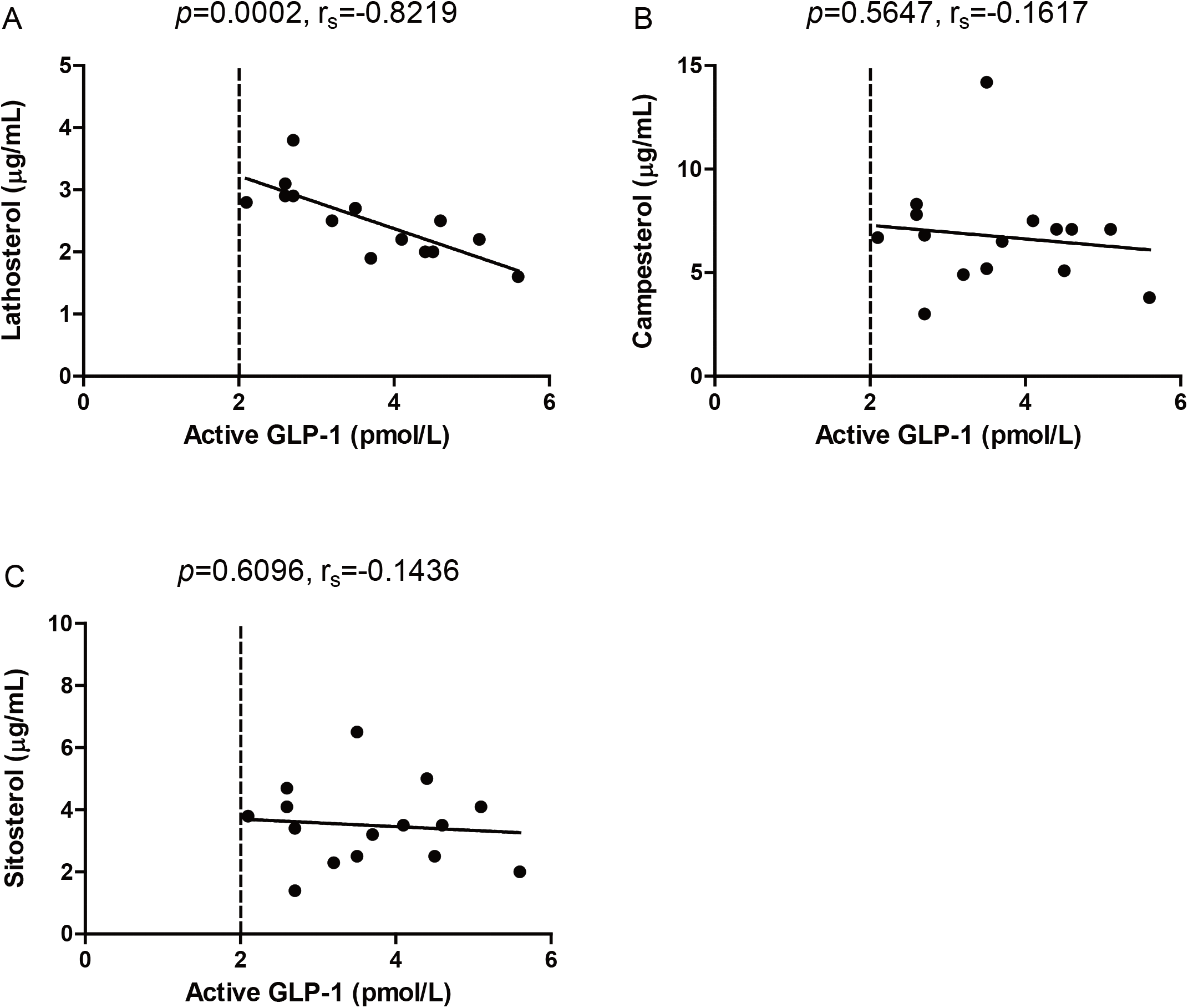
The correlation between fasting active glucagon-like peptide-1 and lathosterol (A), campesterol (B), and sitosterol (C) at the end of this study (n=17) The plots include data on withdrawals. Plots below the detection limit of (2.0pmol/L) plasma GLP-1 were omitted. The figure presents a total of 15 plots.

## 4. Discussion

The results of the present study are consistent with those of previous studies, showing that DPP-4 inhibitors decrease LDL-C levels [6, 7]. Moreover, our results showed that anagliptin significantly decreased fasting plasma lathosterol levels, whereas it did not significantly alter fasting sitosterol or campesterol levels. Thus, anagliptin monotherapy may have reduced cholesterol synthesis in the liver, but it did not appear to have reduced cholesterol absorption in the intestines. Our results suggest that higher fasting active GLP-1 levels are associated with lower fasting plasma lathosterol levels. However, a previous study by Juntti-Berggren et al. [8] showed that continuous administration of GLP-1 to patients with T2DM did not change LDL-C levels. GLP-1 analogs, when injected subcutaneously, directly flows into the systemic circulation. In contrast, active GLP-1, which is elevated by DPP-4 inhibitor, is secreted from L-cells in the ileum but enters the liver via the portal vein before entering the systemic circulation. Therefore, endogenous GLP-1 may exert different effects on cholesterol metabolism, whereas exogenous GLP-1 may not. A previous study has reported that anagliptin administered twice a day increases active GLP-1 levels after dinner significantly more than sitagliptin administered only once a day [4]. Administrating DPP-4 inhibitors twice a day may be more advantageous for lowering LDL-C levels than administering them once a day. Although vildagliptin is also administered twice a day, its half-life is markedly shorter than that of anagliptin, which might lead to different LDL-C lowering effects. Therefore, further studies are warranted. The effect of GLP-1 on 3-hydroxy-3-methyl-glutaryl-CoA (HMG-CoA) activity in the liver is still unclear. In contrast, the administration of anagliptin has been shown to suppress the expression of SREBP-2, a transcription factor for cholesterol synthesis, and decrease LDL-C levels in LDL receptor-knockout mice, a model for studying hyperlipidemia, using that mechanism of action. Thus, the suppression of SREBP-2 by anagliptin appears to be one of the mechanisms underlying the drug’s LDL-C lowering effect [9].

Onoue et al. recently observed that anagliptin, in combination with other hypoglycemic drugs, reduces fasting apoB-48 levels; hence, they proposed that anagliptin is beneficial for lipid metabolism by acting via the inhibition of intestinal lipid transport [10]. This effect was particularly observed in previous studies that included the concomitant use of SUs, which have significant hypolipidemic effects and hypoglycemic activity. It is, therefore, possible that SUs masked the LDL-C-lowering effects of anagliptin, especially in long-term treatments. Another study has showed that anagliptin therapy for 4 weeks decreases the level of the cholesterol synthesis marker lathosterol without affecting the levels of cholesterol absorption markers [11]. The present study differed from these studies with regard to the following: 1) monotherapy was performed for long period of 6 months and 2) the relationship between lathosterol and active GLP-1 levels was examined.

The present results are not consistent with previous findings, which showed that DPP-4 inhibitors decrease post-prandial TG and apoB-48 levels after a fat-loading test in patients with T2DM [12]. Our results showed contradictory results that anagliptin did not decrease post-prandial TG or apoB-48 levels in patients with T2DM after the CLT (Fig. 4A and 4B). As a limitation of the present study, the sample size was small, particularly for determining secondary outcomes. However, the study population was homogeneous for anagliptin monotherapy, thus ruling out any potential influence of antidiabetic and antihyperlipidemic drugs. Another limitation is that the sub-analysis of the correlation between active GLP-1 levels and lathosterol, campesterol or sitosterol includes the data of withdrawals. Therefore, the sub-analysis does not accurately reflect the influence of anagliptin alone.

## 5. Conclusions

Anagliptin monotherapy for 6months significantly decreased the levels of LDL-C and the cholesterol synthesis marker lathosterol without affecting the levels of the cholesterol absorption markers. Lathosterol levels were negatively correlated with fasting active GLP-1 levels.

## Data Availability

The datasets used and/or analyzed during the current study are available from the corresponding author on reasonable request.

## Acknowledgment

The authors thank Ms. Sawako Sato for her technical and secretarial assistance.

